# Machine learning based preoperative analytics for the prediction of anastomotic insufficiency in colorectal surgery: a single-centre pilot study

**DOI:** 10.1101/2021.12.11.21267569

**Authors:** Stephanie Taha-Mehlitz, Larissa Wentzler, Fiorenzo Angehrn, Ahmad Hendie, Vincent Ochs, Victor E. Staartjes, Markus von Flüe, Anas Taha, Daniel Steinemann

## Abstract

**Introduction:** Anastomotic insufficiency (AI) is a relatively common but grave complication after colorectal surgery. This study aims to determine whether AI can be predicted from simple preoperative data using machine learning (ML) algorithms.

**Methods and analysis:** In this retrospective analysis, patients undergoing colorectal surgery with creation of a bowel anastomosis from the University Hospital of Basel were included. Data was split into a training set (80%) and a test set (20%). The group of patients with AI was oversampled to a ratio of 50:50 in the training set and missing values were imputed. Known predictors of AI were included as inputs: age, BMI, smoking status, the Charlson Comorbidity Index, the American Society of Anesthesiologists score, type of operation, indication, haemoglobin and albumin levels, and renal function.

**Results:** Of the 593 included patients, 88 experienced AI. At internal validation on unseen patients from the test set, area under the curve (AUC) was 0.61 (95% confidence interval [CI]: 0.44-0.79), calibration slope was 0.16 (95% CI: −0.06-0.39) and calibration intercept was 0.06 (95% CI: 0.02-0.11). We observed a specificity of 0.67 (95% CI: 0.58-0.76), sensitivity of 0.36 (95% CI: 0.08-0.67), and accuracy of 0.64 (95% CI: 0.55-0.72).

**Conclusion:** By using 10 patient-related risk factors associated with AI, we demonstrate the feasibility of ML-based prediction of AI after colorectal surgery. Nevertheless, it is crucial to include multicenter data and higher sample sizes to develop a robust and generalisable model, which will subsequently allow for deployment of the algorithm in a web-based application.

**Strengths and limitations of this study:**

- To the best of our knowledge, this is the first study to establish a risk prediction model for anastomotic insufficiency in a perioperative setting in colon surgery.
- Data from all patients that underwent colon surgery within 8 years at University Hospital Basel were included.
- We evaluated the feasibility of developing a machine learning model that predicts the outcome by using well-known risk factors for anastomotic insufficiency.
- Although our model showed promising results, it is crucial to validate our findings externally before clinical practice implications are possible.

## Introduction

Anastomotic insufficiency (AI) is a severe complication following gastrointestinal surgery^1^. According to Rahbari et al.^2^, AI is defined as a defect at the anastomotic site which leads to a connection between intra- and extraluminal compartments. AI is itself considered to be an independent risk factor for adverse clinical and oncological outcomes like decreased survival of cancer patients and increased readmission rates after surgery^3–5^. The approximated incidence of AI is 3.3% after colon anastomosis and 8.6% after colorectal anastomosis in specialised centres^4^. However, these rates are likely considerably higher in centres lacking dedicated colorectal surgery teams and after emergency surgery. In fact, depending on diagnostic criteria, AI rates of over 10% have been reported in the literature^6–10^. Hospital stay is extended by twelve days on average, and healthcare-related expenses are increased by up to 30.000 USD in patients who experience AI^1,2^.

In previous publications, a multitude of risk factors for AI has been identified^11,12^. Among these risk factors, age^13^, gender^7,14–19^, body mass index (BMI)^20,21^, Charlson Comorbidity Index (CCI)^14,22^, American Society of Anesthesiologists (ASA) score^13,21–24^, leukocytosis^23^, anaemia^25^, hypoalbuminemia^15,26,27^, steroid use^28,29^, renal function^23^, previous abdominal surgery^16^, active smoking^6,30,31^, and alcohol abuse^6,32^, liver metastasis^16^, indication^16,33^, type of surgery^33^, emergency surgery^34^, surgical approach^35^, anastomotic technique^36,37^, and defunctioning ileostomy^33,35^ were identified more or less consistently. Integrating all of these risk factors into one holistic clinical prediction of AI is a very challenging task, even for experienced physicians. Indeed, even experienced surgeons were reported to systematically underestimate the risk of AI by clinical assessment^38^. Undoubtedly, the ability to preoperatively predict AI precisely would allow for better resource allocation and enhanced patient preparation, and an improved patient-physician relationship due to the improved quality of informed consent.

Machine learning (ML) algorithms can be exceptionally competent at integrating various patient variables into a unified risk model, which is able to generate predictions specifically for each patient. Development and rigorous validation of clinical prediction models require large amounts of multicentre data as well as external validation. Before embarking on said multicentre data collection, piloting a modelling strategy to assess feasibility and to identify the most valuable inputs is crucial. Consequently, this pilot study with data from a single centre aims to assess whether AI can be predicted from simple patient-related risk factors.

## Methods

### Overview and Data collection

Data was extracted retrospectively from the patient registry of the University Hospital of Basel. Patients who underwent colon anastomosis for various reasons including neoplasia, diverticulitis, ischaemia, iatrogenic or traumatic perforation, or inflammatory bowel disease between 1^st^ of January 2012 and 31^st^ of December 2019 were eligible. This study was completed based on the transparent reporting of a multivariable prediction model for individual prognosis or diagnosis (TRIPOD) statement checklist for the development of clinical prediction models^39^. Utilizing the aforementioned data, we developed ML models with the aim of predicting AI, and internally validated the models on held-out test data.

### Ethical Considerations

Ethical approval was provided for this study by the Northwestern and Central Ethics Committee Switzerland (BASEC-Nr 2020-02265).

### Predictors and Outcome Measures

AI was defined according to Gessler et al.^40^ and Rahbari et al.^2^ as any clinical sign of leakage, confirmed by radiological examination, endoscopy, clinical examination of the anastomosis, or upon reoperation.

Recorded variables included risk factors that already have been reported in literature such as age, gender, BMI, active smoking, alcohol abuse (>2 alcoholic beverages per day), prior abdominal surgery, preoperative leucocytosis (≥10.000 per mm^3^), preoperative steroid use, CCI, ASA Score, renal function (CKD Stages G1 to G5), albumin (g/dl), and haemoglobin level (g/dl), liver metastasis, indication, type of surgery, emergency surgery, surgical approach (laparoscopic or open), anastomotic technique (hand-sewn or stapler) and defunctioning ileostomy.

### Model Development

Continuous variables and categorical variables were recorded as mean ± standard deviation (SD). Moreover, categorical variables as decimal numbers instead of percentages. Python 3.6.9 was used to perform all analyses. The Keras library^41^ was used for the artificial neural networks, the XGBoost library^42^ was used for XGBs, and the scikit-learn library^43^ was used for all other architectures.

Data was randomly split into two sets: 80% of the data was used as training data, while the remaining 20% was utilized as a test set to validate the models internally. Initially, the two sets had similar class distribution. However, the training set’s minority class (patients with AI) was oversampled using random oversampling until a 50:50 class distribution was achieved to prevent the models from overpredicting the majority class due to class imbalance^44,45^. Recursive feature elimination was applied to determine the optimum set of relevant features predicting the outcome in the training data set^46^.

A *k*-nearest neighbor (KNN) imputer was co-trained on the pre-oversampling training data to impute any missing data^47^. We trialled multiple model architectures, including generalised linear models (GLMs), support vector machines (SVMs), naïve Bayes classifiers, random forests, extreme gradient boosting machines (XGB), and artificial neural networks. Training occurred in repeated 5-fold cross validation with 10 repetitions to select the optimal model architecture and hyperparameters based on the area under the receiver operating characteristics curve (AUC). Recalibration was applied using Platt’s method^48^ based on the pre-oversampling training set, and the cut-off for binary classification was selected based on AUC on the training set using the closest-to-(0,1)-criterion^49^.

The models were subsequently internally validated using the held-out test set, with ninety-five percent confidence intervals (Cis) being generated using 1000 bootstrap repetitions.

## Results

### Cohort

In the training set, a total of 474 patients were included in the training set, of which 77 (16.2%) suffered from AI. The mean age was 64.6 ± 15.4. Furthermore, 225 (47.5%) patients were male and 249 (52.5%) were female. The mean BMI was 25.1± 5.3 and 152 (32.1%) patients were active smokers. **Table 1** provides an overview of the cohorts.

**TABLE 1.**
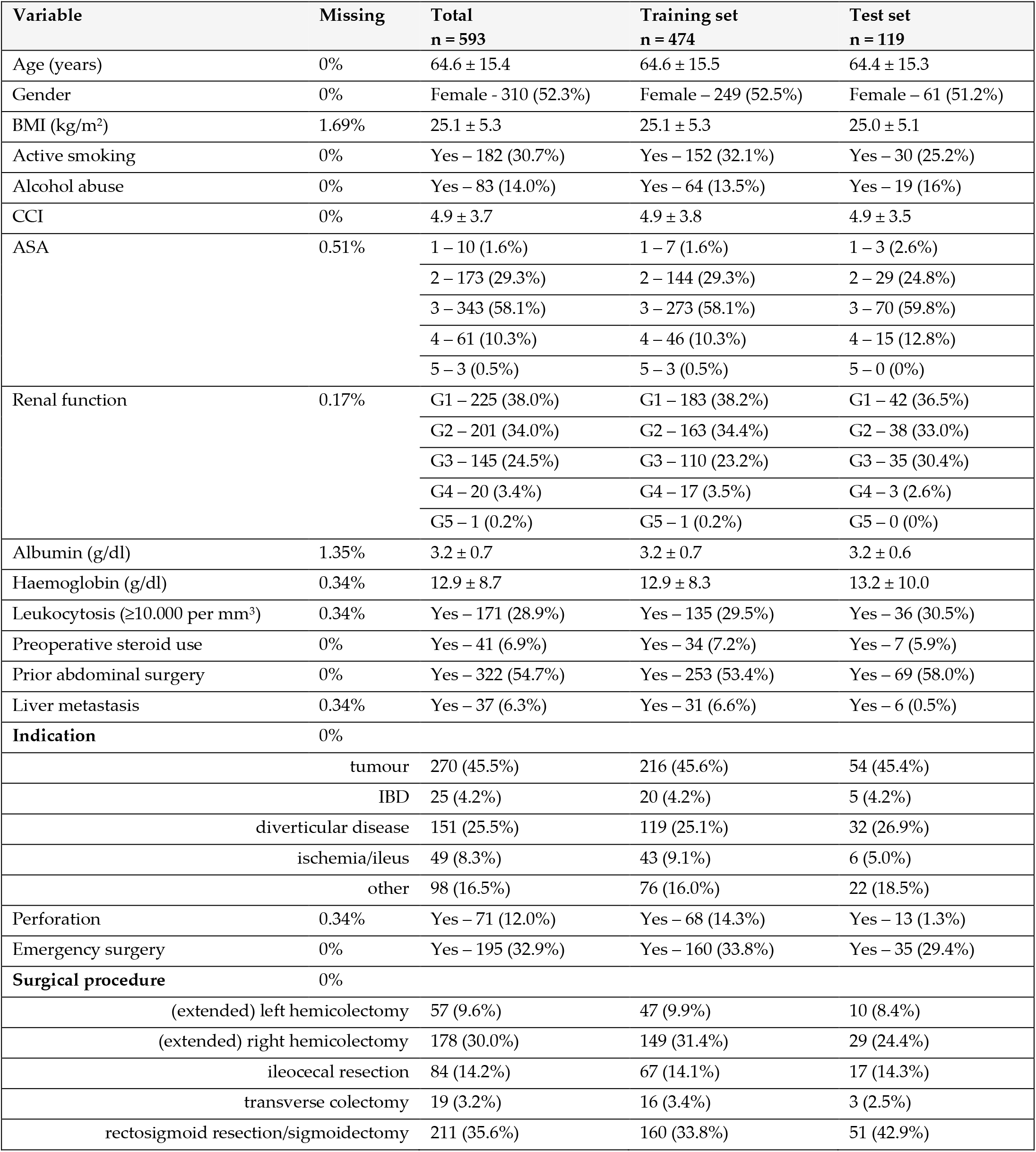
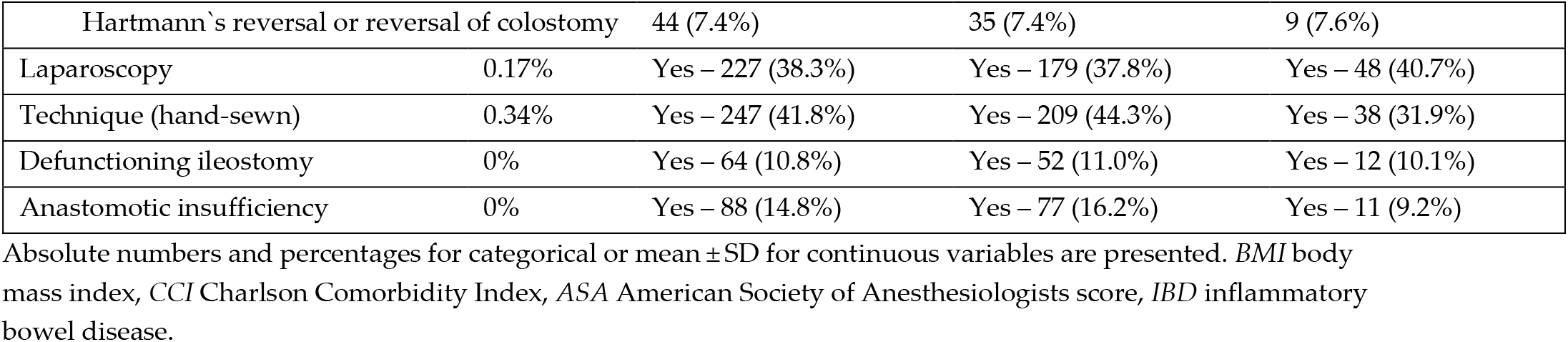
Overview of patient characteristics including missingness.

As for the test set, a total of 119 patients were included, of which 11 (9.2%) suffered from AI. Furthermore, 58 (48.8%) patients were male, and 61 (51.2%) were female. The mean BMI in the test set was 25.0±5.1 and 30 (25.2%) patients were active smokers.

### Model Performance

The performance of all six models was assessed by internal validation using the test data set. This performance is reported in detail along with resampled training performance in **Table 2**. The random forest provided the highest AUC with a value of 0.61 (95% CI: 0.44 – 0.79) while also producing acceptable probabilities based on the calibration slope [0.16 (95% CI: −0.06 – 0.39)], and intercept [0.06 (95% CI: 0.02 - 0.11)]. Moreover, the random forest exhibited a sensitivity of 0.36 (95% CI: 0.08 – 0.67), specificity of 0.67 (95% CI: 0.58 – 0.76) and accuracy of 0.64 (95% CI: 0.55 – 0.72). Specific feature importance within the random forest is displayed in **Table 3**. The calibration and AUC curves for the random forest classifier are displayed in **Figures 1** and **2**.

**TABLE 2.** Performance evaluation of the final random forest classifier. Resampled training performance and test set performance are reported.

| **Metric** | **Training Set** | **Test Set** |
| --- | --- | --- |
| AUC | 0.87 (0.77-0.95) | 0.61 (0.44-0.79) |
| Accuracy | 0.83 (0.76-0.9) | 0.64 (0.55-0.72) |
| Recall | 0.74 (0.53-0.92) | 0.36 (0.08-0.67) |
| Precision | 0.48 (0.3-0.67) | 0.1 (0.02-0.2) |
| Specificity | 0.85 (0.78-0.91) | 0.67 (0.58-0.76) |
| Brier | 0.09 (0.05-0.12) | 0.11 (0.08-0.15) |
| Calibration slope | 1.01(0.74-1.24) | 0.16 (-0.06-0.39) |
| Calibration intercept | -0.01(-0.04-0.03) | 0.06 (0.02-0.11) |

*AUC* area under recall-precision curve

**TABLE 3.**
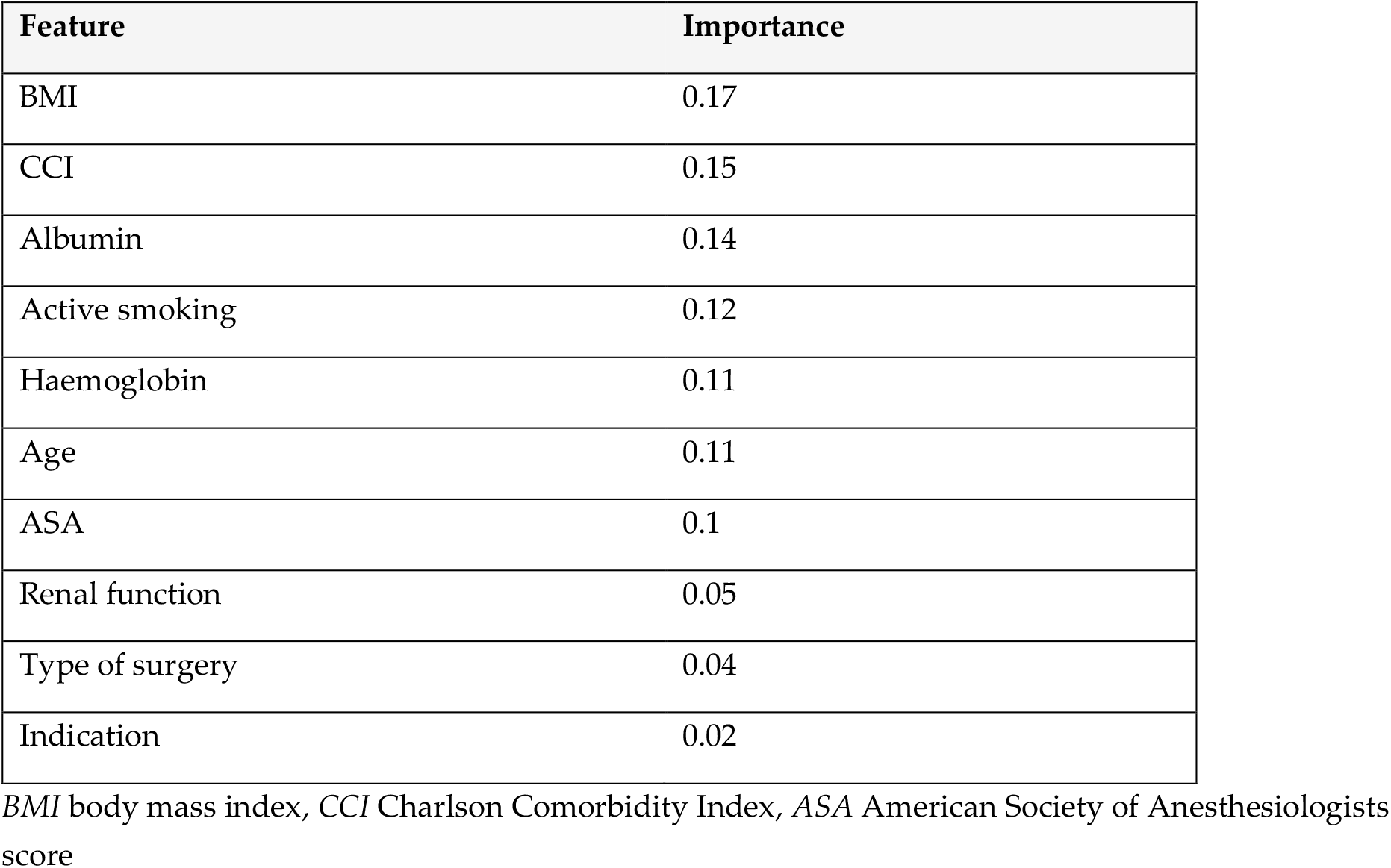
Predictor importance according to random forest classifier.

**FIGURE 1.**
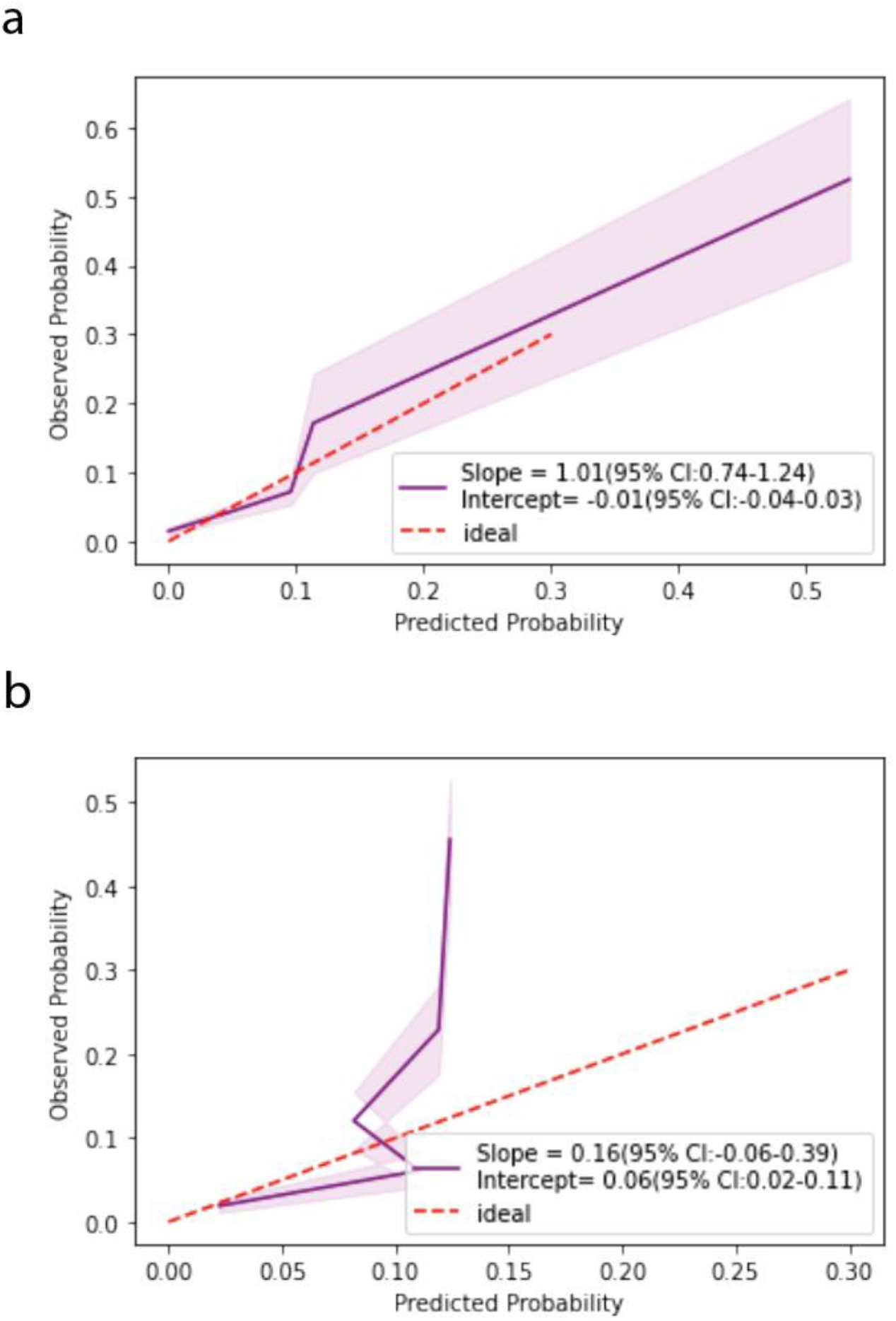
Resampled training (**a**) and test (**b**) set calibration curves of the random forest classifier.

**FIGURE 2.**
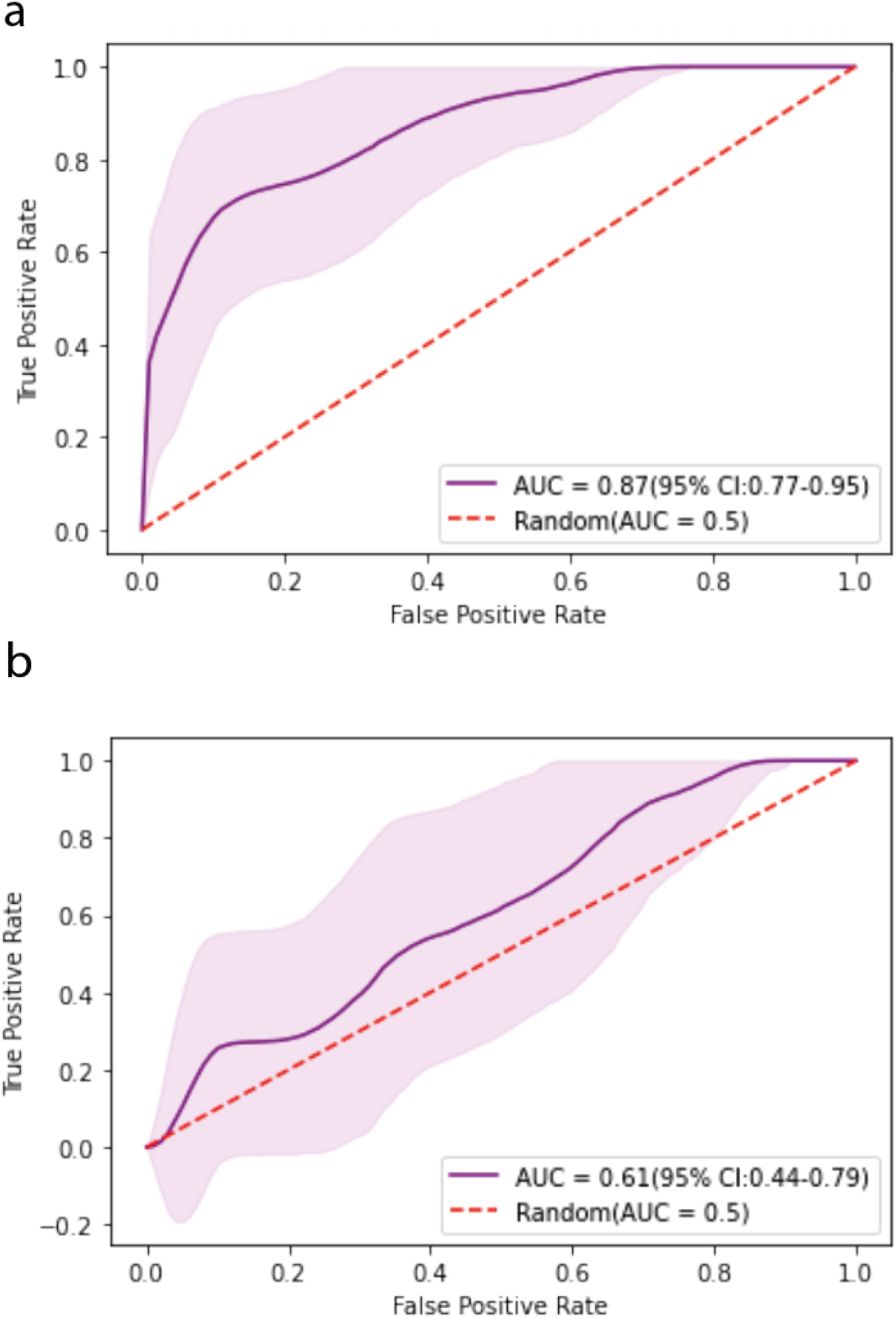
Resampled training (**a**) and test (**b**) set area under the receiver operating characteristics curves (AUC) of the random forest classifier.

## Discussion

In a pilot study using data from a single reference centre for colorectal surgery, we assess the feasibility of predicting AI accurately from simple data using ML techniques. Our findings demonstrate that modelling AI to a certain extent is feasible and identifies the most important input variables, laying the groundwork for more extensive multicentre modelling and external validation.

Even though a plethora of studies have analysed risk factors for AI, up to this day, no reliable clinical prediction model for AI has been established^11–37^. To the best of the authors’ knowledge, this study is the first one attempting to solve the classification problem of AI (‘*will my patient suffer from AI after colorectal surgery*?’) by application of ML algorithms.

There is a widespread misunderstanding that variable importance measures gleaned from clinical prediction models can discover correlations and causalities in the same way that explanatory modelling does (prediction versus explanation)^50^. Indeed, this common misconceptualisation exists because predictive and explanatory modelling are often not as explicitly distinguished as attempted here in this study. Indeed, the interchangeable use of the concepts of *in-sample correlation* and *out-of-sample generalisation* can lead to false clinical decision making^51^. While those variables identified as having high feature importance in this study may indeed be the most crucial ones for precise and generalisable prediction of AI, it cannot safely be concluded that these variables are necessarily also important independent risk factors for AI in their own right.

Another separate question is the initial choice of input variables for clinical prediction modelling, which can be achieved in various ways^52^: In any case, a balance between performance through the inclusion of many variables and between the goal of arriving at parsimonious models that truly generalise needs to be struck. The choice of variables for this study was focused on patient-related risk factors to minimize the statistical noise from differing standard procedures in distinct clinical centres. The following section aims to clarify why the risk factors chosen have not only been correlated to AI previously but are also part of plausible pathomechanisms.

The CCI and the ASA classification system are two commonly used scoring systems assessing the patients’ pre-anaesthesia medical comorbidities and expected mortality risk, respectively. Even though the ability to predict peri- and postoperative complications is claimed for both scores, the ASA score and the CCI give valuable insight into the patients’ general medical conditions and thus can be used to generate more specific scoring systems^47,48^. Indeed, the CCI and ASA score represent the most important features in our final model.

Serum levels of albumin and other hepatic proteins have been established as easily obtainable laboratory parameters for malnourishment^53^. Since malnourishment itself is a risk factor for impaired wound healing in general^54^, it is conceivable that hypoalbuminemia could be factor predisposing patients for AI. In fact, hypoalbuminemia has already been linked to increased postoperative morbidity and mortality in patients with colon cancer^27,55^.

The most important pathophysiological basis for AI is believed to be gastrointestinal ischaemia, decreasing the amount of oxygen available to the recovering intestinal wall^56^. Consequently, the intestinal tissue becomes more dysfunctional, which may lead to delayed wound healing. These suspected pathophysiological processes align well with the factors determined relevant for the prediction of AI risk in our model. In a similar way, haemoglobin levels determine the blood’s capacity to supply peripheral tissues with oxygen^57,58^.

Smoking has been consistently evaluated as a risk factor for AI^15,31^. Furthermore, it is well established that tobacco smoking is among the top risk factors for vascular disease^59^, making it a logical contributor to vascular insufficiency leading to subsequently impaired wound healing and postoperative AI risk.

Our model seems to be more specific than sensitive. This indicates that our model is proficient at ruling in high-risk AI with a positive prediction. Conversely, our model is less precise in ruling out low-risk AI. However, a rule-in model could prove to be of great value for clinicians by simply identifying the very high-risk group. Furthermore, the random forest model seems fairly well-calibrated. Well-calibrated predicted probabilities are arguable more important in clinical practice (“How likely is it that I am going to experience AI?’ – ‘Your probability is 17%.’) instead of binary predictions (‘Am I going to suffer from AI?’ – ‘The model predicts yes/no’). Physicians are experts at dealing with uncertainty and risks, and probabilities are thus more appreciated by patients and physicians than a mere yes or no answer – apart from the fact that patients are never binary but instead represent a spectrum of risk^60^.

Another difficulty in clinical prediction modelling is choosing the appropriate sample size. According to a common rule of thumb, there should be at least 10 minority class observations in a dataset per feature^61^. This study relies on 10 patient-related risk factors, thus making a total number of 1000 patients with AI who would be necessary for training, while we finally were able to include 88 patients with AI. Other architectures such as random forests, artificial neural networks, and SVMs seem to require much more data per feature^62^. Therefore, it is conceivable that including more patients might further refine the model. Consequently, it is crucial to recruit more patient data from other hospitals in a multicentre study.

Clinical prediction models can facilitate assessing individual risks and make more informed decisions based on predictive analytics that are tailored to each patient. However, especially in colorectal surgery, the indication for surgery is only rarely truly elective. Therefore, a prediction model can only help decide whether an intervention should be postponed to improve the risk profile or, especially for emergency interventions, whether a patient would benefit from a diverting stoma to minimize and modify risk factors before re-joining the colon. On the other hand, a comprehensive predictive model may also increase a patient’s acceptance of the primary placement of a protective stoma. Thus, such a model could potentially also help to improve the physician-patient relationship through enhanced patient education.

The results of a predictive model cannot be seen as a clear recommendation pro or contra an intervention as the risk profile it mirrors is tailored only to a specific endpoint and thus does not entirely reflect the patient’s global situation. Indeed, components of decision-making such as the psychological distress of a patient with chronic diverticulitis are not included in the model and have a decisive influence on the indication. Consequently, prediction models should be seen only as adjunctive information to be used in a complementary way for informed shared decision-making. Nevertheless, the necessity for evidence-based clinical prediction models becomes clear when considering the relative inability of even experienced clinicians in predicting clinical outcomes^38^, while the ethical implications of an ‘AI doctor’ technology independent from human control have to be taken into account, too^63^. Consequently, ML-based clinical prediction models could be deemed a contemporary optimal trade-off between the clinical experience of human experts and the exploitation of big data by learning algorithms.

### Limitations

The relatively high AI rate of 14.8% in our cohort can be seen as a limitation. Similarly, the difference in AI incidence among training versus test set represents an additional hurdle that is realistic, as AI rate is described inconsistently in the literature^4,6–10^. The patient population at the University Hospital Basel with 32.9% emergency cases and a cohort that includes transplanted and immunosuppressed patients is expected to have higher complication rates^34^. Nevertheless, such a difference to other hospitals should be reflected in the ASA score, the CCI, and blood values and thus also in our results. By including patient data from other institutions in future analyses, this number will be balanced out, and a differentiated breakdown according to emergency interventions, immunosuppressant use, previous radio-/chemotherapy, and cancer diagnosis, which additionally reflect a patient’s health status, is conceivable and could be implemented in our ML-algorithm.

One further caveat of any model is the danger of overfitting. In the context of clinical prediction modelling, overfitting means that an algorithm adheres too strictly to the training data, especially its inherent variance and possible noise factors (e.g. noise generated by a hospital’s standardized procedures). With enough training, the algorithm will perform extremely well on the training data while losing its generalisation capability towards new data from other centres. Indeed, it is not unlikely that this study might suffer from slight overfitting due to standardized hospital procedures because the data was exclusively collected in one clinical centre. However, this weakness could be addressed by recruiting more patient data from other hospitals. Lastly, we have purposefully not yet deployed our model for clinical application in, e.g. a web-app, as any single-centre and not yet externally validated clinical prediction model is not yet recommended for clinical use^64^.

## Conclusion

In this pilot study, by using 10 patient-related risk factors associated with AI, we demonstrate the feasibility of ML-based prediction of AI after colorectal surgery. Nevertheless, it is crucial to include multicentre data and higher sample sizes to develop a robust and generalisable model, which will subsequently allow for deployment of the algorithm in a web-based application.

## Data Availability

All data produced in the present study are available upon reasonable request to the authors.

## Author Contributions

Conceptualization A.T., V.S.; data collection L.W., F.A., S.T-M.; administration and ethics S.T.-M., A.T., F.A., D.S.; analysis A.H., V.S., A.T., V.O.; visualization A.H., S.T.-M.; writing—original draft preparation A.T., S.T.-M., V.O., A.H.; writing—review and editing V.S., F.A., D.S., M.v.F.; All authors have read and agreed to the published version of the manuscript.

